# Sleep disturbance as a precursor to anxiety, depression, and PTSD among rural Kenyans: a cross-lagged panel analysis from a rural Kenyan interventional cohort

**DOI:** 10.1101/2023.11.11.23298315

**Authors:** Michael L. Goodman, Miryoung Lee, Andrew Springer, Vanessa Schick, Elizabeth Vaughan, Christine Markham, Stanley Gitari, Fridah Mukiri

## Abstract

Sleep quality is essential to biopsychosocial functioning, yet there remains limited longitudinal research on sleep and mental or social well-being within low- or middle-income countries. This study utilizes longitudinal cohort data from a community-based empowerment program in Meru County, Kenya to assess cross-lagged correlations between sleep disturbance, social support, symptoms of depression, anxiety, and posttraumatic stress,

Participants (n=373; 92% women; age range 18-86 years) who reported more sleep disturbance at T1 reported significantly more symptoms of depression, anxiety and PTSD, and significantly less social support at T2 (average 11 weeks later), controlling for all within-time correlations across measures, within-measure correlations across time, and sociodemographic background characteristics.

Findings are consistent with research across high-income countries, underscoring the need for more contextualized research into sleep behaviors across low- and middle-income countries. Findings may inform interventions to increase mental and social well-being within Kenya.

## Introduction

Quality sleep, marked by adequate continuity, duration, and frequency, is essential to human well-being. Sleep health is multidimensional, adapted to contextual considerations, and characterized by subjective satisfaction, appropriate timing, adequate duration, high efficiency, and sustained alertness during waking hours (Buysse, 2014). Sleep disturbance, defined as sleep that is inadequate in duration or quality, is correlated with impaired functioning of multiple physiological systems (Orr, Fass, Sundaram, & Scheimann, 2020; Chong et al., 2022). Sleep disturbance and related sleep disorders have been found to correlate with negative mental, behavioral, and physical health outcomes across all age groups (Ohayon, Carskadon, Guilleminault, & Vitiello, 2004). Investigators have identified many affective, cognitive, attentional, and motivational correlates of and sequelae to sleep disturbance including post-traumatic stress disorder (PTSD), depression and anxiety (Zhai, Zhang, & Zhang, 2015; Hertenstein et al., 2019; Freeman et al., 2020; Shalev, Liberzon, & Marmar, 2017).

Sleep disturbance is considered a component feature of PTSD, anxiety, and depression (Shalev, Liberson, & Marmar, 2017; Chellappa & Aeschbach, 2022). Investigators have sought to disentangle the temporal relations between sleep disturbance and mental health challenges. Multiple prospective studies in higher-income countries have demonstrated that sleep disturbance, most commonly insomnia, may predict higher incidence and symptomology of subsequent depression, anxiety, and PTSD (Zhai, Zhang, & Zhang, 2015; Cox & Olatunji, 2016). Many cross-sectional studies have been analyzed in attempts to show causality between poor mental health and sleep disturbance (Oh et al., 2019; Du et al., 2020), potentially contributing to confusion regarding the temporal order of measured variables (MacKinnon, Krull, & Lockwood, 2000). A 2021 meta-analysis of randomized control trials in high-income countries show improved sleep quality causally leads to improve mental health – including symptoms of depression and anxiety, and PTSD (Scott et al., 2021).

In addition to mental health sequalae of sleep disturbance, poor sleep correlates with impairment to many social processes (Gordon, Carrillo, & Barnes, 2021). Within mostly cross-sectional data, social correlates of poor sleep include increased physiological sensitivity to social-evaluative tasks, worse identification of social cues and social isolation, relationship strain, intimate partner violence victimization, and smaller social networks (Gordon, Carrillo, & Barnes, 2021). While there are few longitudinal studies assessing potential bidirectionality between social relationships and poor sleep quality, poor sleep has been shown to predict feelings of anger and worsened perceived relationship quality (Audigier, Glass, Slotter, & Pantesco, 2023).

Nationally representative data from the United States demonstrate that sleep disturbance is more common among people who are socio-economically marginalized and engage less in healthy behaviors (e.g. unhealthy diet patterns, limited exercise, use of cigarettes and alcohol), raising concerns for health equity in promoting sleep quality (Hale, Troxel, & Buysse, 2020).

Given the wide-ranging correlates and consequences of sleep disturbance and emerging concerns for health equity in promoting quality sleep, the global health agenda should include sleep quality as a potentially modifiable determinant of global population health (Chattu et al., 2019). Yet, the overwhelming evidence on sleep quality and health has originated from high-income countries and China (Chattu et al., 2019; Cosgrove et al., 2020). Extant research has identified associations between sleep quality and health correlates within low- or middle-income countries that parallel those found in high-income countries (e.g. Stranges et al., 2012). However, research that has been conducted on sleep in low- and middle-income countries has been almost entirely cross-sectional or limited to select clinic populations (Stranges et al., 2012; Simonelli et al., 2018; Wang, Cheng & Xu, 2019). The dearth of longitudinal data on sleep quality, mental health, and social relationships from within sub-Saharan Africa, and other low- or middle-income regions has unfortunately led to continual reliance on findings from western cultures to drive global mental health program and policy planning (Herman et al., 2022).

Global health researchers, organizations, and activists have increasingly sounded alarm regarding the real global increase in common mental disorders (Herman et al., 2022). Social support is imperative for mental health, and promotes resilience to various threats to ecological, social, mental, and physical health – further underscoring the need to understand potential determinants of social support (Surkalim et al., 2022; Harandi, Taghinasab, & Nayeri, 2017). Clarifying relationships between sleep disturbance, social support, and mental well-being is essential to promoting global health and development.

The estimated burden of mental health disorders within Kenya, and globally, is increasing (GBD 2019 Mental Disorders Collaborators, 2022). There is a need for a “whole of society” approach to improving mental health across Kenya and other low- or middle-income countries (Herrman et al., 2022). Given the demonstrated impact of sleep disturbance on mental health within high-income countries, it is essential to understand how sleep disturbance may impact the population mental health within Kenya.

### Study Aim

This study seeks to identify temporal relationships between sleep disturbance, social support and common mental disorder symptoms (i.e., symptoms of depression, generalized anxiety, and PTSD) among adults in semi-rural Kenya across two waves of an interventional cohort. Briefly, the intention behind the study intervention was to support the nurturing capacities of families and communities to which street-involved children are reintegrating and to support primary prevention of street-migration of children (Goodman et al., 2023).

## Methods

### Study design and participant selection

This study analyzes longitudinal interventional cohort data. All participants in the program are 18 years of age or older – though this is not an interventional requirement. The intervention is an active program with open enrollment, and no comparisons are made in this study between intervention participants and non-participants.

Participants in this study were selected at random from among participants in a multi-sectoral, multi-level, community-based empowerment program in Meru County, Kenya. The intervention design has been described more completely previously (Goodman et al., 2021). Initial participants are recruited to the program through identification as a known family member of a child living on the streets (Goodman et al., 2023). Index families – the first within a village to join the program – invite neighbors to form an internal microlending group comprised of 25-30 participating families. Subsequent recruitment to the program within a village follows word-of-mouth and is open to anyone interested in joining. While all participants in the present study were intervention participants at the times of data collection, not all participants were “index families” – that is, most participants did not have a child living on the streets at the time of the study.

After 4-months of establishing group-based microfinance practices, lending groups are invited to nominate one or two participants to form a Parliamentary Committee to reflect the interests of and advocate on behalf of participants. We deliver a novel, 6-month curriculum integrating positive psychology, psychological flexibility, interpersonal theory, and strengths-based growth. Upon completing this curriculum, participants are invited to form Resource Committees reflecting shared interests – e.g. HIV prevention, positive parenting, business development, etc.

Previous research on the program model indicates depression decreases with time in the program, potentially mediated by decreased loneliness and increased access to social resources (Goodman et al., 2021). Additionally, access to social resources available through the intervention moderates associations between prior adversity and future generalized anxiety among participants in the program (Goodman et al., 2022).

### Random selection process

With the aim of recruiting a representative sample of adult participants from participants, recruitment from the program to this interview questionnaire was conducted by offering each willing participant the chance to select a piece of paper from an opaque bag. The opaque bag contained folded pieces of paper, with “1” representing being selected to this study and “0” representing not being selected to this study. On the first interview day, seven pieces of paper with the number “1” were included in the opaque bag, and an additional “0” was added until there was an even number of willing participants and pieces of paper. Interviews were delivered in the location where the groups met, removed from the remaining participants.

Selected survey participants joined the program in late 2022, and completed standardized survey questionnaires in February or March 2023 (T1), and again in April or May 2023 (T2).

### Inclusion criteria

Inclusion criteria required study participants to be active members of an internal lending group, present at the weekly session from which data were collected, 18 years or older, and willing to engage in the interview.

### Survey conduct

Each survey was delivered by a local, trained, paid language expert – most of whom were nursing students at a nearby university or recent graduates.

### Measures

Survey questionnaires utilized previously validated psychometric scales. Primary measures in this analysis included sleep disturbance, depression, anxiety, PTSD, and social support. All primary measures were analyzed as continuous variables. Local language experts worked in separate teams to translate questionnaires into the local language, Kimeru, and back to English. Comparisons between initial and back-translated items were conducted by these two teams, and an additional native English speaker (MG). Items in conflict were resolved by agreement between these teams, and administered to a small Kimeru-speaking sample (n=32) to assess psychometric properties. Items on all psychometric scales were averaged to contribute to a summative measure, following inspection of factor structure and internal reliability.

Sleep disturbance was measured using the 8-item PROMIS sleep disturbance scale (Cronbach’s alpha (α)=0.84 at T1; α=0.84 at T2; Yu et al., 2012). The scale asks respondents to recall their sleep over the past 7-days and respond to various statements about the quality and quantity of their sleep such as, “My sleep was restless” using a 5-point Likert-type response. The PROMIS sleep disturbance scale previously showed associations with higher prevalence of PTSD following road traffic accidents in an Ethiopia population (Fekadu et al., 2019).

Depression was measured using the 21-item Beck’s Depression Inventory-II (α=0.81 at T1; α=0.84 at T2; Beck, Steer & Brown, 1996). The BDI-II asks participants to reflect on their current thoughts and feelings covering multiple domains of depression – including negative or flattened affect, loss of appetite, and reduced motivation – on a 4-point response format. The BDI-II has been used and validated in diverse global populations (e.g. Win, Kawakami, & Htet Doe, 2019).

Generalized anxiety disorder was measured using the GAD-7 (α=0.86 at T1; α=0.89 at T2; Spitzer, Kroenke, Williams, & Lowe, 2006). The GAD-7 is a 7-item anxiety scale using a 2-week recall period, prompting respondents to report the frequency of experiencing nervousness, worry, difficulty relaxing, and irritability. The GAD-7 has previously been validated in Kenya and other global populations (Goodman et al., 2022; Dhira, Rahman, Sarker, & Mehareen, 2021).

PTSD was measured using the Primary Care PTSD Screen for DSM-5 (PC-PTSD-5) (Prins et al., 2016). While developed to screen for PTSD among veterans, the PC-PTSD-5 index has been shown to have excellent diagnostic accuracy among civilian populations (Williamson et al., 2021). Within the present sample, the PC-PTSD-5 showed high internal reliability (Kuder-Richardson (KR)-20=0.76 at T1; KR-20=0.8 at T2). The PC-PTSD-5 screens respondents for the presence of any life-threatening experience over the respondent’s lifetime. Country-specific population-based estimates of lifetime exposure to such events range globally from 29% to 83% (Benjet et al., 2016). Respondents who indicate the presence of such an experience are then asked 5-items documenting the presence or absence of PTSD symptoms (e.g. experiential avoidance, related nightmares, being easily startled, experiencing guilt or self-blame, feeling numb or detached) over the previous month. The PC-PTSD-5 has previously been adapted for use in South Africa, Zimbabwe, and Uganda, and found to be associated with forced sex, and risk-taking in these contexts (Webb et al., 2022). Respondents who reported no prior life-threatening traumatic event, or who reported no symptoms from such an event, were considered “0” within this analysis.

Social support was measured using the brief 2-way social support scale (SSS) (Obst, Shakespeare-Finch, Krosch, & Rogers, 2019). The brief 2-way SSS assesses the giving and receiving of emotional and instrumental support in the present, with 12 items such as “there is at least one person that I can share most things with” and “I give others a sense of comfort in times of need” (α=0.86 at T1; α=0.89 at T2). Within multiple cultural contexts, this relatively new measure of social support is associated with mental well-being as expected (Liu et al., 2022).

Control variables. To control for within-subject variations potentially associated with sleep behaviors, we included measures of wealth, income, marital status, age, and years of formal schooling. Wealth index was recorded as household asset ownership of 12 common items (KR-20: 0.72). Income was recorded as a continuous variable reflecting estimated household monthly income. Marital status was recorded as a binary variable – married / living with someone as though married vs. not married / not living with someone. Age was recorded in years since birth. Formal schooling was measured as an ordinal variable between no school “0” and finishing a tertiary program “14.”

### Statistical Analysis

Descriptive analyses that included univariate mean (SD) and pairwise Spearman rank sum correlations were conducted of all primary and control variables. The correlation matrix included Bonferroni-adjusted significance levels (α=0.05).

Wilcoxon matched-pairs signed-rank tests were used to assess the statistical equivalence of all continuous measures between T1 and T2.

The primary analysis for this study utilized cross-lagged panel analysis, with a full set of potential pathways between each of the 5 primary, continuous variables (sleep disturbance, depressive symptoms, anxiety symptoms, PTSD symptoms, and social support) at T1 and T2. This model was created using structural equation modeling using a backward modeling process, removing correlations until only correlations under the alpha threshold (α<0.2) remained in the model. Control variables were added to all pathways, and further removed until only correlations under the alpha threshold (α<0.2) remained. Correlations between variables at T1 and T2 were included, and removed if under the alpha threshold (α<0.2). To be considered significant, path coefficients must be under a final alpha threshold (α<0.05). SEM correlation standard errors were calculated using robust variance estimators to account for variables with non-normal distributions (Mansournia et al., 2021).

Additionally, we presented the standardized SEM coefficients between control and primary variables at T2 to enhance understanding of these relationships. All analyses were conducted using Stata v.16 (StataCorp, 2019).

### Ethical consideration

Data were collected following ethical approval from the Institutional Review Boards at the Kenya Methodist University and the University of Texas Medical Branch in Galveston, TX. All participants provided informed written consent prior that emphasized the voluntary and confidential nature of the study to participating in the study. Compensation ($1) was given to participants’ microfinance group on behalf of each study participant, following a practice designed by community members interested in ensuring equitable distribution of available resources following a random selection.

## Results

The mean (SD) for each variable is shown in Table 1. Eighty-seven percent (87%) of respondents reported experiencing a life-threatening experience during their lifetime (T1). Symptoms of depression, anxiety, and PTSD significantly decreased from T1 to T2 (p≤0.01). Social support did not change significantly between the two waves. The mean (sd) age was 41.2 years (13 years). Respondents owned, on average, 5.4 items on the 12-item wealth index, which did not significantly change between the two time points. Mean monthly household income (USD) significantly increased from $34 (T1) to $45 (T2). Mean (sd) years of formal education was 5.2 (3.2), and 75% of respondents were married or living with a partner as though married. The mean (sd) number of weeks between two waves was 11 (0.9).

**Table 1.** Descriptive characteristics of sample.

|  | n | T1 |  | T2 |  | p-value |
| --- | --- | --- | --- | --- | --- | --- |
|  |  | Mean | ±SD | Mean | ±SD |  |
| Depression (range: 0-63) | 373 | 8.1 | 7.2 | 6.5 | 6 | **<0.001** |
| Anxiety (range 0-21) | 362 | 22.4 | 6 | 20.2 | 15.9 | **<0.01** |
| PTSD: Report previous life-threatening experience (T1) | 368 | 87% | 0.3 |  |  |  |
| PTSD (range 0-5) | 368 | 3 | 1.7 | 2.7 | 1.9 | **0.01** |
| Sleep disturbance (range: 0-4) | 364 | 1.23 | 0.97 | 1.23 | 0.9 | *Ns* |
| Social support (range: 1-6) | 363 | 4.9 | 0.8 | 4.9 | 0.9 | *Ns* |
| Age (years) | 373 | 41.2 | 13 | 41.5 | 13 | *Ns* |
| Wealth index | 373 | 5.4 | 2.6 | 5.5 | 2.6 | *Ns* |
| Income (USD per month) | 357<sup>t</sup> | 33.5 | 34 | 44.9 | 27.2 | **<0.001** |
| Education (years) | 373 | 5.2 | 3.2 | 5.3 | 3.1 | *Ns* |
| Female gender | 373 | 92% | 0.3 |  |  |  |
| Married / Living w partner (yes) | 373 | 76% | 0.4 | 75% | 0.4 | *Ns* |
| Weeks between observations | 361 |  |  | 11.3 | 0.9 |  |

Notes: Descriptive characteristics of sample. All tests of independence within continuous variables were calculated using the Sign Rank test and $\chi^2$ tests were used for categorical comparisons. Due to floor effects on Sleep disturbance (T1), we provide bivariate comparisons between T1 and T2 Sleep disturbance among respondents who report any sleep disturbance at T1 - 85% of the sample.

Table 2 demonstrates the correlation matrix with significance between primary and control measures. Significant pairwise correlations exist between sleep disturbance (T1) and symptoms of depression (T1; r=0.35, p<0.001; and T2, r=0.29, p<0.001), and anxiety (T1, r=0.21, p<0.01; and T2, r=0.24, p<0.001). Sleep disturbance (T2) is significantly correlated with symptoms of depression (T2, r=0.38, p<0.001), symptoms of anxiety (T2, r=0.29, p<0.001), and social support (T2, r=-0.32, p<0.001).

**TABLE 2:**
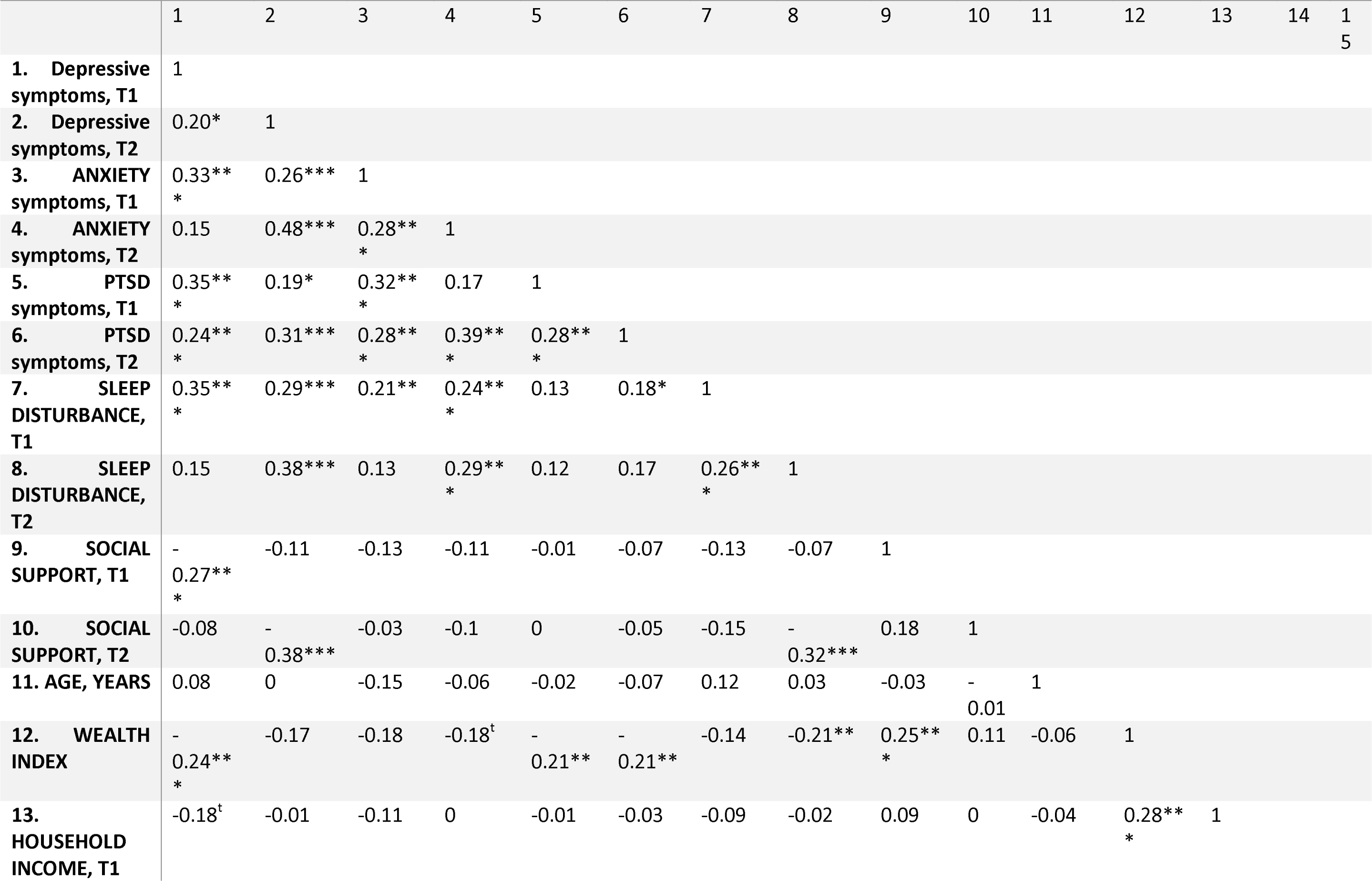
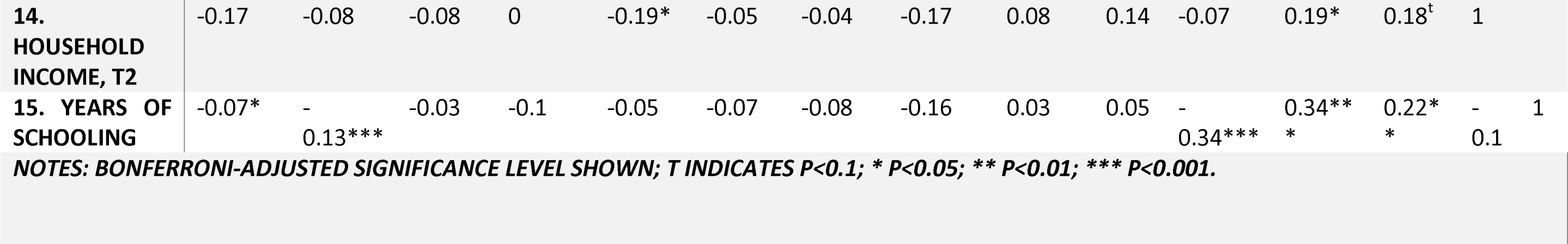
SPEARMAN CORRELATION MATRIX OF PRIMARY AND CONTROL MEASURES.

The cross-lagged panel analysis, displayed in Figure 1 below, indicates that more sleep disturbance at T1 predicts subsequently worse symptoms of depression (T2; r=0.25; p<0.001), anxiety (r = 0.19; p<0.001), PTSD (T2; r=0.14; p=0.01), and less social support (T2; r= -0.14; p<0.01) after controlling for within-variable and within-time correlations between primary measures.

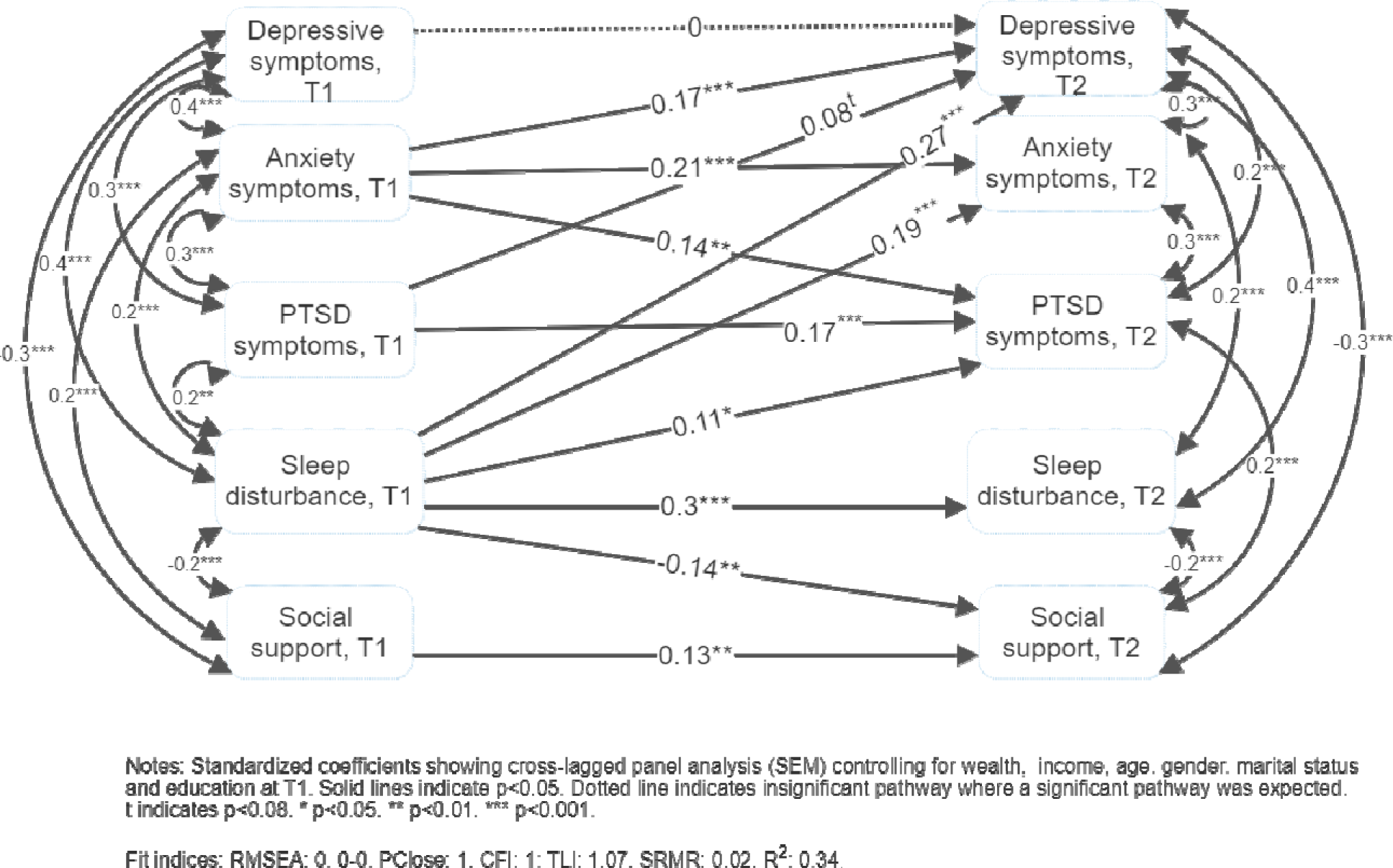

More depressive symptoms (T2) were better predicted by prior symptoms of anxiety (T1; r =0.19; p<0.001) and sleep disturbance (T1; r=0.25; p<0.001) than by previous depressive symptoms (T1, r=0; p=0.9). Anxiety symptoms (T1) also predicted symptoms of PTSD (T2; r=0.16, p<0.001) and later anxiety symptoms (T2, r=0.21; p<0.001).

Social support was significantly correlated with fewer symptoms of depression, anxiety and sleep disturbance at T1, and with fewer symptoms of depression, PTSD and less sleep disturbance at T2. Social support was not predictive of any included variable.

Table 3 shows the standardized correlation coefficients between included control variables and primary variables (T2). As shown, more years of schooling (reported at T1) predicted less sleep disturbance at T2 (r=-0.1; p<0.05), but no other control variable was statistically significant. Married respondents reported significantly higher social support (T2; r=0.12; p<0.05) and fewer symptoms of depression (T2; r=-0.12; p=0.01). Wealth predicted significantly fewer symptoms of PTSD (T2; r=-0.13; p<0.05).

**Table 3:** SEM-based cross-lagged panel analysis with control variables.

| T2 variable | T1 variable | $\beta$ | 95% CI | |
| --- | --- | --- | --- | --- |
| Sleep disturbance, T2 | Sleep disturbance, T1 | 0.24*** | 0.14 | 0.34 |
|  | Years of schooling | -0.11* | -0.2 | -0.01 |
|  | Married, T1 | -0.06 | -0.16 | 0.04 |
| Social support, T2 | Sleep disturbance, T1 | -0.14** | -0.24 | -0.04 |
|  | Social support, T1 | 0.13** | 0.03 | 0.23 |
|  | Married, T1 | 0.13** | 0.02 | 0.24 |
| Depressive symptoms, T2 | Sleep disturbance, T1 | 0.25*** | 0.15 | 0.35 |
|  | Depressive symptoms, T1 | 0 | -0.1 | 0.1 |
|  | Anxiety symptoms, T1 | 0.19*** | 0.10 | 0.27 |
|  | PTSD symptoms, T1 | 0.08<sup>t</sup> | 0 | 0.16 |
|  | Married, T1 | -0.12* | -0.21 | -0.02 |
| Anxiety symptoms, T2 | Sleep disturbance, T1 | 0.19*** | 0.09 | 0.28 |
|  | Anxiety symptoms, T1 | 0.21*** | 0.11 | 0.26 |
|  | Age, years, T1 | -0.07<sup>t</sup> | -0.21 | 0 |
| PTSD symptoms, T2 | Sleep disturbance, T1 | 0.14** | 0.04 | 0.24 |
|  | Anxiety symptoms, T1 | 0.16** | 0.07 | 0.26 |
|  | PTSD symptoms, T1 | 0.19*** | 0.09 | 0.29 |
|  | Wealth, T1 | -0.13* | -0.23 | -0.02 |
|  | Female gender, T1 | 0.08 | -0.02 | 0.17 |
|  | Age, years, T1 | -0.06 | -0.15 | 0.04 |

Notes: Primary and control variables (T1) correlated with primary variables (T2) from final SEM model. <sup>t</sup> indicates $p < 0.1$ ; \* $p < 0.05$ ; \*\* $p < 0.01$ ; \*\*\* $p < 0.001$ . Robust variance estimators used to calculate standard errors.

## Discussion

This novel study investigated temporal relationships between sleep disturbance, mental health and social well-being in a general population within Meru County, Kenya. Similar to patterns identified elsewhere (e.g. Franzen & Buysse, 2022; Hertenstein et al., 2019; Freeman et al., 2020; Shalev, Liberzon, & Marmar, 2017; Audigier et al., 2023), sleep disturbance predicted more symptoms of depression, anxiety, PTSD, and less social support after 11 weeks of follow-up.

By contrast, and unexpected by us, sleep disturbance was not predicted by any previous primary measure (i.e., anxiety symptoms, depressive symptoms, or social support) when we adjusted for socioeconomic status variables. These findings suggest that sleep quality is fundamental to mental health and social functioning serving as a precedent rather than an outcome. Our study findings suggest that sleep merits much greater focus by global mental health researchers, advocates and interventionists than it previously has received. Despite the importance of sleep for mental health and overall health-as found in this study and others, there are few sleep clinics within Kenya or other sub-Saharan countries, and existing sleep clinics mostly respond to patients with the capacity to receive a diagnosis and the financial means to access specialty clinics (Komolafe et al., 2021). Given the fundamental salience of sleep function to mental and social health and previous econometric evaluation of lost productivity due to poor sleep (Van Stolk, 2017), greater investment in understanding sleep processes at a population-level within the Kenyan context is required.

Unfortunately, we were not able to identify many variables that predicted sleep disturbance in this study, preventing meaningful nuance to subsequent investigations. Higher education attainment was found to predict less subsequent sleep disturbance, though income, wealth, age, marital status, and gender were not. Bivariate correlation analysis between control variables at T1 and sleep disturbance at T2 provides little information that might otherwise nuance these null findings from the full SEM model. Most bivariate relations between control variables (T1) and sleep disturbance (T2) were also non-significant.

More basic behavioral research is required to understand the cultural and contextual factors contributing to, or potentially improving, sleep disturbance. Prior research in higher-income countries has found stress management, relaxation practice, stimulus control, sleep hygiene, and physical exercise may improve sleep distrubance (Murawski et al., 2018). Urban living has previously shown to be a greater risk for sedentary lifestyles – one common predictor of poor sleep within higher income countries (Martins et al., 2021; Yang, Shin, Li, & An, 2017). In the rural study context, where only 13% of respondents reporting owning motorized transportation (item #12 in the wealth index), sedentary behaviors are less likely to contribute to poor sleep in our study. Pain has been identified as contributing to poor sleep, and was unmeasured within the study data (Finan, Goodin, & Smith, 2013).

Sleep disturbance as a predictor of reduced social support is an important finding. Social support and social functioning are essential to human flourishing and a central element within the Flourishing Community interventional model from which these data were collected (Goodman et al., 2023). This finding suggests the need for a deeper understanding of the processes by which social support is developed and maintained within the interventional context and the mechanisms by which sleep disturbance may both undermine, and be alleviated by, these processes. Economic modelling, comparing productive work time between individuals with different durations of average nightly sleep, finds that poor sleep contributes to a 1%-3% reduction in potential national economic productivity in five Organization for Economic Cooperation and Development (OECD) countries (Hafner, Stapanek, Taylor, Troxel, & Van Stolk, 2017). Whether sleep contributes to economic functioning merits research within low- and middle-income countries, and may be mediated by worse psychosocial functioning.

While we expected temporal relationships between sleep disturbance and poor mental health, we did not anticipate depressive symptoms (T1) would be less predictive of (T2) depressive symptoms than were symptoms of anxiety, PTSD, or sleep disturbance. This finding suggests, perhaps, that persistent depressive symptoms are mediated by mental and behavioral processes better registered by the sleep-, anxiety-, and PTSD-related measures. Future research should add more waves of measurement, and qualitative investigation with people experiencing persistent depression to better understand operative mechanisms, and potential interventional points to reduce persistent depression. At minimum, these findings suggest improving sleep quality among participants may improve subsequent mental health – potentially interrupting otherwise persistent mental health challenges.

The percentage of respondents reporting any experience of life-threatening trauma over the course of their lives was slightly higher than national prevalence estimates in countries like the United States or South Africa (87% within sample vs. 83% within the United States; 74% in South Africa; Benjet et al., 2015). We find these differences credible, as the sample population was selected due to identification of street-involved children migrating from those villages – and socio-ecological risk factors of street-migration of children may well increase the trauma risk for other individuals within the same villages. Future studies should adopt or adapt a life events checklist to improve the specificity of life-threatening traumatic histories (e.g. Kwobah et al., 2022). Trauma research may reveal population-level nuances regarding stress responses to traumatic events, the role of sleep in protecting against stress responses, and socio-cultural factors potentially providing resilience to diverse traumatic events.

We interpreted responses to validated psychometric scales in many epidemiologic studies as reflecting symptomatic severity rather than diagnosed mental illness, as there are limited trained clinical diagnosticians within the study location – and across Kenya, sub-Saharan Africa and the world (Keynejad, Spagnolo, & Thornicroft, 2021). Thus, while this study is informed by practical constraints within the context, these constraints are found globally. To address the need for improved understanding and the development of public health approaches to community mental health therapeutics,the National Institutes of Health has led the development of an alternative to diagnostic-based approaches to mental health that primarily serve the clinician-patient relationship found within the Diagnostic and Statistical Manual (DSM-V) or ICD-10 diagnostic coding (Cuthbert, 2022). The NIH Research Domain of Criteria (RDoC) posits sleep quality as one essential component of mental and behavioral health (Kozak & Cuthbert, 2016). Our findings support the intent of the RDoC program by showing more sleep disturbance predict relatively worse future mental health symptomologies and less social support – and provide a route to exploring mechanisms to improve population-level mental and social well-being by reducing sleep disturbance. There is increasing recognition for the need to move beyond diagnostic criteria to support individual clinical encounters and towards a public health-enabled approach to mental health, which is consistent with the RDoC program and our aims in this study (Cosgrove et al., 2020).

Finally, these data were not collected from a random sample of the population but rather randomly from an intervention program open to all members of the population. Future studies should clarify differences between study participants and the broader population with reference to interventional effectiveness and differences informing the decision to participate in the program. It is possible, and probable, that baseline sleep disturbance moderates any effectiveness of the intervention on mental and social well-being. Clarifying whether and how sleep disturbance moderates interventional effectiveness should be an essential task in global mental health promotion.

### Limitations

This analysis relies on interviewer-administered self-reported data; using interviewers was necessary to include participants with low or no functional literacy (15% of respondents reported never attending school). Self-reported data are subject to reporting biases such as social desirability bias and recall bias. Respondents, influenced by social desirability bias, would need to indicate worse than average sleep but average or better mental and social well-being at T1 and average sleep but worse than average mental and social well-being at T2 for social desirability bias to contribute to observed correlations. We think it is unlikely social desirability bias would lead to this differential reporting between time points. If social desirability bias leads to differential reporting between time points, it seems just as likely that misclassification could occur in the opposite direction (that is: over-reported sleep quality relative to mental and social health at T1 and under-reported sleep quality relative to mental and social health at T2). We believe findings are robust to influence by social desirability bias, though assert they are not evidence of causation – only of temporal relations controlling for other between- and within-panel correlations. Future research should investigate whether sleep quality is causally related to subsequent mental health as found in other contexts; research in any context is required to establish causal relationships between sleep and social variables.

A review comparing reliability of objective measures of sleep to self-reported sleep quality found comparable reliability is lower than one may expect (Cudney et al., 2022). The extent to which available objective measures of sleep relate temporally or causally to mental, social, or physiological factors within Kenya, sub-Saharan Africa, and globally warrants more research. Within these data, we find significant and predictable relationships between subjective sleep quality, reflecting a 7-day recall period, and subsequently worse mental and social well-being. Beyond considering other measures of sleep quality – including objective observations, daily journal, and other psychometrically valid measures – future research should consider other physiological, behavioral, social and economic outcomes and contexts (cf. Cudney et al., 2022 for review). Ultimately, findings indicate a potentially substantial, and under-identified, contribution of sleep to mental and social well-being within Kenya.

The generalizability of findings is limited by the over-representation by women within the sample. Future studies should investigate temporal relationships between sleep disturbance and psychosocial functioning across durations of time, as this study was limited to a 3 month observation period.

## Conclusion

Sleep is a fundamentally necessary element of human health, impacting biophysiological processes, mental health and social functioning. We present a novel, temporal analysis of sleep disturbance and subsequently worse symptoms of depression, anxiety, PTSD and lower social support. Findings suggest that sleep is a fundamental determinant of mental and social well-being within this context, though research to generalize findings is required. Findings indicate the strong need for further research investment on sleep characteristics, determinants and consequences across populations.

## Data Availability

All data produced in the present study are available upon reasonable request to the authors

## References

Audigier, A., Glass, S., Slotter, E. B., & Pantesco, E. (2023). Tired, angry, and unhappy with us: Poor sleep quality predicts increased anger and worsened perceptions of relationship quality. Journal of Social and Personal Relationships, 10.1177/02654075231193449.

Beck, A. T., Steer, R. A., & Brown, G. (1996). Beck depression inventory–II. Psychological assessment.

Benjet, C., Bromet, E., Karam, E. G., Kessler, R. C., McLaughlin, K. A., Ruscio, A. M., & Koenen, K. C. (2016). The epidemiology of traumatic event exposure worldwide: results from the World Mental Health Survey Consortium. Psychological medicine, 46(2), 327–343.

Chattu, V. K., Manzar, M. D., Kumary, S., Burman, D., Spence, D. W., & Pandi-Perumal, S. R. (2019). The global problem of insufficient sleep and its serious public health implications. Healthcare, 7(1). 10.3390/healthcare7010001

Chellappa, S. L., & Aeschbach, D. (2022). Sleep and anxiety: From mechanisms to interventions. Sleep medicine reviews, 61, 101583.

Chong, P. L., Garic, D., Shen, M. D., Lundgaard, I., & Schwichtenberg, A. J. (2022). Sleep, cerebrospinal fluid, and the glymphatic system: A systematic review. Sleep medicine reviews, 61, 101572.

Cosgrove, L., Mills, C., Karter, J. M., Mehta, A., & Kalathil, J. (2020). A critical review of the Lancet Commission on global mental health and sustainable development: Time for a paradigm change. Critical Public Health, 30(5), 624–631.

Cox, R. C., & Olatunji, B. O. (2016). A systematic review of sleep disturbance in anxiety and related disorders. Journal of anxiety disorders, 37, 104–129.

Cudney, L. E., Frey, B. N., McCabe, R. E., & Green, S. M. (2022). Investigating the relationship between objective measures of sleep and self-report sleep quality in healthy adults: a review. Journal of Clinical Sleep Medicine, 18(3), 927–936.

Cuthbert, B. N. (2022). Research domain criteria (RDoC): progress and potential. Current Directions in Psychological Science, 31(2), 107–114.

Dhira, T. A., Rahman, M. A., Sarker, A. R., & Mehareen, J. (2021). Validity and reliability of the Generalized Anxiety Disorder-7 (GAD-7) among university students of Bangladesh. PLoS one, 16(12), e0261590.

Du, C., Zan, M. C. H., Cho, M. J., Fenton, J. I., Hsiao, P. Y., Hsiao, R., & Tucker, R. M. (2020). Increased resilience weakens the relationship between perceived stress and anxiety on sleep quality: a moderated mediation analysis of higher education students from 7 countries. Clocks & sleep, 2(3), 334–353.

Finan, P. H., Goodin, B. R., & Smith, M. T. (2013). The association of sleep and pain: an update and a path forward. The journal of pain, 14(12), 1539–1552.

Freeman, D., Sheaves, B., Waite, F., Harvey, A. G., & Harrison, P. J. (2020). Sleep disturbance and psychiatric disorders. The Lancet Psychiatry, 7(7), 628–637.

GBD 2019 Mental Disorders Collaborators. (2022). Global, regional, and national burden of 12 mental disorders in 204 countries and territories, 1990–2019: a systematic analysis for the Global Burden of Disease Study 2019. The Lancet Psychiatry, 9(2), 137–150.

Goodman, M. L., Baker, L., Maigallo, A. K., Elliott, A., Keiser, P., & Raimer-Goodman, L. (2022). Adverse childhood experiences, adult anxiety and social capital among women in rural Kenya. Journal of Anxiety Disorders, 91, 102614

Goodman, M. L., Elliott, A. J., Gitari, S., Keiser, P., Onwuegbuchu, E., Michael, N., & Seidel, S. (2021). Come together to decrease depression: Women’s mental health, social capital, and participation in a Kenyan combined microfinance program. International Journal of Social Psychiatry, 67(6), 613–621.

Goodman, M. L., Elliott, A. J., Gitari, S., Keiser, P., Raimer-Goodman, L., & Seidel, S. E. (2021). Come together to promote health: case study and theoretical perspectives from a Kenyan community-based program. Health Promotion International, 36(6), 1765–1774.

Goodman, M., Seidel, S. E., Springer, A. E., Elliott, A., Markham, C. M., Serag, H., & Gitari, S. (2023) Enabling structural resilience of street-involved children and youth in Kenya: reintegration outcomes and the Flourishing Community model. Frontiers in Psychology, 14, 1175593.

Gordon, A. M., Carrillo, B., & Barnes, C. M. (2021). Sleep and social relationships in healthy populations: A systematic review. Sleep Medicine Reviews, 57, 101428.

Hafner, M., Stepanek, M., Taylor, J., Troxel, W. M., & Van Stolk, C. (2017). Why sleep matters—the economic costs of insufficient sleep: a cross-country comparative analysis. Rand health quarterly, 6(4), 11. PMID: 28983434.

Hale, L., Troxel, W., & Buysse, D. J. (2020). Sleep health: an opportunity for public health to address health equity. Annual review of public health, 41, 81–99.

Harandi, T. F., Taghinasab, M. M., & Nayeri, T. D. (2017). The correlation of social support with mental health: A meta-analysis. Electronic physician, 9(9), 5212.

Herrman, H., Patel, V., Kieling, C., Berk, M., Buchweitz, C., Cuijpers, P., & Wolpert, M. (2022). Time for united action on depression: a Lancet–World Psychiatric Association Commission. The Lancet, 399(10328), 957–1022.

Hertenstein, E., Feige, B., Gmeiner, T., Kienzler, C., Spiegelhalder, K., Johann, A., & Baglioni, C. (2019). Insomnia as a predictor of mental disorders: a systematic review and meta-analysis. Sleep medicine reviews, 43, 96–105.

Keynejad, R., Spagnolo, J., & Thornicroft, G. (2021). WHO mental health gap action programme (mhGAP) intervention guide: updated systematic review on evidence and impact. BMJ Ment Health, 24(3), 124–130.,

Komolafe, M. A., Sanusi, A. A., Idowu, A. O., Balogun, S. A., Olorunmonteni, O. E., Adebowale, A. A., & Mosaku, K. S. (2021). Sleep medicine in Africa: past, present, and future. Journal of Clinical Sleep Medicine, 17(6), 1317–1321.

Kozak, M. J., & Cuthbert, B. N. (2016). The NIMH research domain criteria initiative: background, issues, and pragmatics. Psychophysiology, 53(3), 286–297.

Kwobah, E. K., Misra, S., Ametaj, A. A., Stevenson, A., Stroud, R. E., Koenen, K. C., & Atwoli, L. (2022). Traumatic experiences assessed with the life events checklist for Kenyan adults. Journal of affective disorders, 303, 161–167.

Liu, T. W., Ng, S. S., Tsoh, J., Chen, P., Xu, R. H., Wong, T. W., & Tse, M. M. (2022). Translation and initial validation of the Chinese (Cantonese) brief 2-way social support scale for use in people with chronic stroke. BioMed Research International, 2022, 10.1155/2022/3511631.

MacKinnon, D. P., Krull, J. L., & Lockwood, C. M. (2000). Equivalence of the mediation, confounding and suppression effect. Prevention science, 1, 173–181.

Mansournia, M. A., Nazemipour, M., Naimi, A. I., Collins, G. S., & Campbell, M. J. (2021). Reflection on modern methods: demystifying robust standard errors for epidemiologists. International Journal of Epidemiology, 50(1), 346–351.

Martins, L. C. G., Lopes, M. V. D. O., Diniz, C. M., & Guedes, N. G. (2021). The factors related to a sedentary lifestyle: A meta-analysis review. Journal of advanced nursing, 77(3), 1188–1205.

Murawski, B., Wade, L., Plotnikoff, R. C., Lubans, D. R., & Duncan, M. J. (2018). A systematic review and meta-analysis of cognitive and behavioral interventions to improve sleep health in adults without sleep disorders. Sleep medicine reviews, 40, 160–169.

Obst, P., Shakespeare-Finch, J., Krosch, D. J., & Rogers, E. J. (2019). Reliability and validity of the Brief 2-Way Social Support Scale: an investigation of social support in promoting older adult well-being. SAGE open medicine, 7, 2050312119836020.

Oh, C. M., Kim, H. Y., Na, H. K., Cho, K. H., & Chu, M. K. (2019). The effect of anxiety and depression on sleep quality of individuals with high risk for insomnia: a population-based study. Frontiers in neurology, 10, 849.

Ohayon, M. M., Carskadon, M. A., Guilleminault, C., & Vitiello, M. V. (2004). Meta-analysis of quantitative sleep parameters from childhood to old age in healthy individuals: developing normative sleep values across the human lifespan. Sleep, 27(7), 1255–1273.

Orr, W. C., Fass, R., Sundaram, S. S., & Scheimann, A. O. (2020). The effect of sleep on gastrointestinal functioning in common digestive diseases. The Lancet Gastroenterology & Hepatology, 5(6), 616–624.

Prins, A., Bovin, M. J., Smolenski, D. J., Marx, B. P., Kimerling, R., Jenkins-Guarnieri, M. A., & Tiet, Q. Q. (2016). The primary care PTSD screen for DSM-5 (PC-PTSD-5): development and evaluation within a veteran primary care sample. Journal of general internal medicine, 31(10), 1206–1211.

Scott, A. J., Webb, T. L., Martyn-St James, M., Rowse, G., & Weich, S. (2021). Improving sleep quality leads to better mental health: A meta-analysis of randomised controlled trials. Sleep medicine reviews, 60, 101556.

Shalev, A., Liberzon, I., & Marmar, C. (2017). Post-traumatic stress disorder. New England journal of medicine, 376(25), 2459–2469.

Simonelli, G., Marshall, N. S., Grillakis, A., Miller, C. B., Hoyos, C. M., & Glozier, N. (2018). Sleep health epidemiology in low and middle-income countries: a systematic review and meta-analysis of the prevalence of poor sleep quality and sleep duration. Sleep Health, 4(3), 239–250.

Spitzer, R. L., Kroenke, K., Williams, J. B., & Löwe, B. (2006). A brief measure for assessing generalized anxiety disorder: the GAD-7. Archives of internal medicine, 166(10), 1092–1097.

StataCorp. 2019. Stata Statistical Software: Release 16. College Station, TX: StataCorp LLC.

Stranges, S., Tigbe, W., Gómez-Olivé, F. X., Thorogood, M., & Kandala, N. B. (2012). Sleep problems: an emerging global epidemic? Findings from the INDEPTH WHO-SAGE study among more than 40,000 older adults from 8 countries across Africa and Asia. Sleep, 35(8), 1173–1181.

Surkalim, D. L., Luo, M., Eres, R., Gebel, K., van Buskirk, J., Bauman, A., & Ding, D. (2022). The prevalence of loneliness across 113 countries: systematic review and meta-analysis. Bmj, 376.

Wang, X., Cheng, S., & Xu, H. (2019). Systematic review and meta-analysis of the relationship between sleep disorders and suicidal behaviour in patients with depression. BMC psychiatry, 19(1), 1–13.

Webb, E. L., Dietrich, J. J., Ssemata, A. S., Nematadzira, T. G., Hornschuh, S., Kakande, A., & Fox, J. (2022). Symptoms of post-traumatic stress and associations with sexual behaviour and PrEP preferences among young people in South Africa, Uganda and Zimbabwe. BMC Infectious Diseases, 22(1), 1–11.

Win, K. L., Kawakami, N., & Htet Doe, G. (2019). Factor structure and diagnostic efficiency of the Myanmar version BDI-II among substance users. Annals of general psychiatry, 18, 1–7.

Yang, Y., Shin, J. C., Li, D., & An, R. (2017). Sedentary behavior and sleep problems: a systematic review and meta-analysis. International journal of behavioral medicine, 24, 481–492.

Yu, L., Buysse, D. J., Germain, A., Moul, D. E., Stover, A., Dodds, N. E., & Pilkonis, P. A. (2012). Development of short forms from the PROMIS™ sleep disturbance and sleep-related impairment item banks. Behavioral sleep medicine, 10(1), 6–24.

Zhai, L., Zhang, H., & Zhang, D. (2015). Sleep duration and depression among adults: A meta-analysis of prospective studies. Depression and anxiety, 32(9), 664–670.

